# Retrospective on Maternal Mortality: Birth Preparedness Among Women At Antenatal Care Clinics in Makeni City, Sierra Leone

**DOI:** 10.64898/2026.06.11.26355446

**Authors:** Tamba Matturie, Abraham Isiaka Jimmy, Tenneh Millicent Conteh, Alusine Rhoda Thullah, Magdaline Philip Umoh, Rebecca Esliker, Lee Presley Gary

## Abstract

Birth preparedness is a critical strategy aimed at promoting safe childbirth by encouraging pregnant women and their families to create thoughtful birth plans and prepare for potential complications. This approach ensures timely access to skilled maternity and health care services, which are essential for reducing maternal mortality. This study assessed the factors influencing birth preparedness among pregnant women attending Antenatal Care Clinics at the Regional Referral Hospital in Makeni City, Sierra Leone. A probability sampling method was used to select 112 pregnant women, and data were collected during 2023 with a structured questionnaire, using the Matturie Birth Preparedness Scale, as uniquely designed and prepared for this study. The collected data were analyzed using STATA software (version 14.0). Our findings revealed significant gaps in birth preparedness: 83.0% of respondents were unaware of their expected delivery date, 79.5 % did not register for antenatal care in their first trimester, and 72.3% had not chosen a delivery location. A striking 92.9% had not identified a potential blood donor. Knowledge gaps were evident, with 62.5% lacking childbirth knowledge and 55.4% unaware of pregnancy complications. Overall, only 17.86%(= 0) of respondents were genuinely prepared for childbirth. Our study highlights a significant lack of birth preparedness among pregnant women in Makeni City, Sierra Leone, with low awareness of critical factors such as expected delivery dates, danger signs, and prenatal emergency planning.

## 1. INTRODUCTION

Our research explored the factors affecting birth preparedness among pregnant women attending an Antenatal Care Clinics (ANC) in the Makama section of the City of Makeni, Bombali District, Northeast Province, Sierra Leone, West Africa. This research was conducted in the Regional Referral Hospital in Makama. In the era of the Millennium Development Goals (MDGs) in 2015, according to estimates, there are 216 maternal deaths per 100,000 live births worldwide [8]. Almost all of these deaths occurred in developing countries, including Sierra Leone. Sometimes, pregnant women and their neonates, most especially in the sub Saharan region of Africa where health service is challenging, lose their lives or encounter morbidity which they will live with for the rest of their lives. But making deliberate plans before, during and after pregnancy will help to achieve a safer motherhood, thereby promoting the health of the mother and the child.

Birth Preparedness (BP) is a plan or measures put in place to reduce delays whenever the need for healthcare service arises and requires access to the requisite services [4]. In both developed and developing countries, the plague called Maternal Mortality (MM) and morbidity still ravage the globe that killed approximately 295,000 women in 2017 alone of which lack of birth preparedness is seen to be one of the contributing factors.

Maternal mortality (MM) is defined as a situation wherein a woman dies while pregnant or when giving birth to a child or 42 days after she has been delivered. It is important to note that maternal mortality can be prevented according to the report from the World Health Organization (WHO) for 2017 which shows an average of 810 women died from causes that can be avoided or prevented every day. According to the report, sub-Sahara Africa (SSA) and Southern Asia (SA) accounted for 86% of the deaths which is 254,000 of the maternal mortality that occurred globally. Of this number, Sub-Sahara Africa (SSA) recorded 2/3 which is 196,000 maternal deaths [13]. The Sierra Leone Demographic Health and Survey SLDHS (2013) showed the country’s maternal mortality ratio (MMR) to be 1,165 per 100,000 live births, making Sierra Leone the highest in the world and six times higher than the global average. Thus, maternal deaths account for 36% of all deaths of women aged 15-49 in Sierra Leone. Also, Sierra Leone Maternal Death and Surveillance Report (MDSR, 2016). SLDHS (2013) points out that Sierra Leone has one of the highest estimated maternal mortality rates in the world at 803 deaths per 100,000 live births. In the UK it is just 9 deaths per 100,000 live births. Even in neighboring Ghana, the figure is only 319 per 100,000 live births. The sad truth is that most of these maternal deaths are preventable if proper care is available and provided. According to the Borgen Project report in 2020, Sierra Leone was amongst the three countries with the highest maternal mortality ratio of 1,120 deaths per 100.000 live births. As the world is moving towards bringing maternal mortality ratio below 10 shows that the updated medical knowledge and current technology to decrease maternal mortality to almost zero is achievable. One of the strategies to apply is to improve health infrastructure and health education in resource constrain settings or developing countries. The lack of access to health facilities and medical professionals is one of the fundamental reasons for maternal mortality; Adolescent pregnancy is another key reason for maternal death especially in the sub-Saharan Africa [6]. In Sierra Leone, maternal deaths accounts for 36% of deaths among women within the ages of 15 – 49 years.

Since free healthcare was introduced in Sierra Leone, there has been remarkable improvement for maternal, neonates and child health,and skilled birth delivery increased from 61% in 2013 to 93% in 2017 and most of these deliveries arenow carried out in a healthcare facility. However, this improvement has not created positive health outcomes when considering maternal mortality records. In the 2017 maternal death review and surveillance report, it was clearly indicated that 79% of the maternal deaths took place within the health facility. Going through such reports for a country like Sierra Leone, one will be left with no option but to try to understand what are the responsible factors. In low income countries like Sierra Leone, income generation for childbirth has always been a huge challenge wherein pregnant women or their families will not be able to seek appropriate healthcare that they can afford and, as a result, a high level of mortality or morbidity may occur. Cultural and behavioral factors are also another challenge; some cultural practices as a result of tradition also have some negative impact on one’s health. Socio-demographic factors like age, religion, marital status and level of education. Birth preparedness are a safe motherhood strategy that aims to promote the timely use of skilled birth attendants and babies assistance during childbirth or obstetric emergencies due to delays. Birth preparedness is one of the effective ways of reducing maternal mortality because it is simple, practicable and result-oriented, but women are not using it, maybe as a result of some other contributing factors [1].

Birth preparedness is a plan that a woman makes by putting in place certain modalities which can be important to her when thinking of becoming a mother. This plan has practices to embark on prior to pregnancy and during pregnancy what to do and what not to do like identifying a place to be delivered in time of labor, means of transportation to access the identified health facility, saving money for delivery expenses, identifying a birth attendant, who makes or take decisions in case of any complication during delivery, a companion or someone who takes care of her while in the hospital [2, 7]. Moreover, a study conducted by the United Nation Development Programme (UNDP), Sierra Leone Annual Report in 2015; Sierra Leone has one of the world’s highest maternal mortality rates, with 1360 deaths per 100,000 live births [9]. More research establishes a theory that birth preparedness reduces the delays in obtaining emergency care [5]. Therefore, this study seeks to assess the factors that affect birth preparedness (BP) among pregnant women attending Antenatal Care Clinics in the Regional Referral Hospital in Makeni City.

## 2. STUDY METHODS and MATERIALS

### 2.1 Study Area

Makeni City is a cosmopolitan center with commercial and administrative activities carried out by the inhabitants; and the main sources of livelihood are farming and trading. It is the largest city in the Northern Province of Sierra Leone. The Regional Referral Hospital in the Northern Province of the country is in Makeni City, situated in the Makama Community, where data for this research study were collected. All the participants were pregnant women who regularly attend Antenatal Care Clinics in the Makeni Regional Referral Hospital.. Every participant had to be a pregnant woman that was attending the antenatal clinics in the Regional Referral Hospital in Makeni.

### 2.2 Study Design, Participants and Sampling Techniques

Our research team used a cross-sectional study design with analysis by type. The cross-sectional design was used to describe the factors influencing birth preparedness among pregnant women in the Northern Province of Sierra Leone. A cross-sectional study is an example of an observational study design. In a cross-sectional study, the researcher looks at data at a particular point in time. The participants are only chosen based on the inclusion and exclusion criteria established for the investigation. Cross-sectional designs are used for population-based surveys and provide a 1-time measurement of the prevailing characteristics of a population. A cross-sectional study can swiftly examine any potential relationships at a particular period in time, despite its inability to demonstrate cause and effect.

### 2.3 Sample Size Determination

The Yamane formula was used for attaining the sample size for the quantitative part of this study (95% confidence level and 0.09 Margin of error). The formula for calculating the sample size is given by:

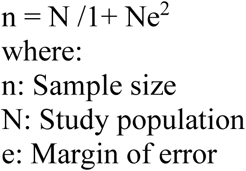

Example Calculation For a study population N = 732 and a margin of error e = 0.09, the sample size is calculated as follows:

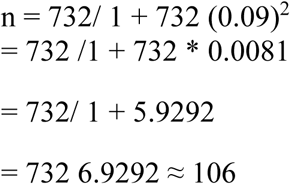

Addition of 6% non-response rate, ≈ 6 participants. Therefore at 95% confidence level and 0.09 precision rate, the sample size for this study was 112. Probability sampling technique was used for this study. Using probability proportion to size sampling is an approach that is useful when the units are of unequal sizes, and ensures the likelihood of a unit being selected is proportionate to the size of the represented population.

### 2.4 Data Collection Tools

Data collection for this was attained with the use of structured questionnaire and for the birth preparedness component, the researcher developed a scale that was used as a guide for screening for birth preparedness, which was referred to as the Matturie birth preparedness scale (M10)score guide). Below is the outline of the score guide:

**Table 1:** Birth Preparedness (Using Matturie Birth Preparedness Scale-M10)

|  | Please tick Yes, or No for the following indicators | Yes (score 1) | No(score 0) |
| --- | --- | --- | --- |
| 1 | Do you know your expected date of delivery? |  |  |
| 2 | Have ever been registered for antenatal care within the first trimester? |  |  |
| 3 | Have you been able to identify birth place? |  |  |
| 4 | During this pregnancy have you identified skilled birth attendant? |  |  |
| 5 | Have you done your shopping for birth supplies for delivery? |  |  |
| 6 | Have you identified a means of transport in case of emergency? |  |  |
| 7 | Have you got a person to accompanying you during an emergency? |  |  |
| 8 | Have you identified a predetermined place to access emergency services? |  |  |
| 9 | Do you have someone to donate blood for you if needed? |  |  |
| 10 | Have you been saving money in case of an emergency? |  |  |

#### Interpretation of Scores

Our researcher developed the following scale to be used as a guide for screening for birth preparedness. The options are outlined below:

M10 Score of 10: Likelihood of being prepared for birth (delivery),

M1o Score of 0-4 = Likely Not prepared for birth (delivery)

M10 Score of 5-10 =Likely prepared for birth (delivery

Research assistants were trained on how to administer the questionnaire and data collection procedures. Successive study participants were identified by the principal researcher following the inclusion and exclusion criteria. Following the selection of the responders, each study participant was given a personalized explanation of the study in a language they could understand preferably Krio.

### 2.5 Data Analysis

For this research, the analysis was carried-out in line with the objectives of the research, a statistical package called STATA was used and a logistic regression analysis was carried out for contingent valuation process. The result was then presented in tables and for percentages, frequencies and proportion it was presented in figures and also in other form such as graph. The odds ratios (ORs), coefficient (Coef) with a 95% confidence interval (CIs) were all calculated to measure the outcome. These statistical measures provide a clear overview of the central tendencies and distributions within the dataset, aiding in the interpretation of key findings.

## 3. RESULTS

### 3.1. Demographic Characteristics of Respondents

The study provides a detailed socio-demographic profile of the participants, who were all pregnant women attending Antenatal Care Clinics at the Regional Referral Hospital in Makeni City during the research period. The participants varied in age, with a significant portion being 20 years and younger, followed by those aged 26–30, 31–35, 21–25, and a smaller group above 35 years. The participants were nearly evenly split between Christians and Muslims. A majority were single, followed by married women, while a small number were divorced, and an even smaller proportion were widowed. Education levels among the participants were mixed: a notable portion had no formal education, some had primary education, a larger group had secondary education, and others had attended tertiary institutions.

**Table 2:** Socio-demographic characteristics of respondents.

| Variable | Frequency (n=112) | Percentage (%) |
| --- | --- | --- |
| **Age category of respondents** |  |  |
| 20 and Below | 47 | 41.96 |
| 21-25 | 13 | 11.61 |
| 26-30 | 25 | 22.32 |
| 31-35 | 16 | 14.29 |
| Above 35 | 11 | 9.82 |
| **Religion of Respondents** |  |  |
| Christianity | 53 | 47.32 |
| Muslim | 59 | 52.68 |
| **Marital status of Respondents** |  |  |
| Divorce | 3 | 2.68 |
| Married | 48 | 42.86 |
| Single | 60 | 53.57 |
| Widow | 1 | 0.89 |

| **Level of Education** |  |  |
| --- | --- | --- |
| Non-formal | 29 | 25.89 |
| Primary | 9 | 8.04 |
| Secondary | 46 | 41.07 |
| Tertiary | 28 | 25 |

**Source: field data (2022)**

### 3.2. Level of Birth Preparedness and Knowledge Level in Pregnancy Danger Signs

The table 4.2 shows the level of birth preparedness of the respondents (pregnant women) who attended antenatal clinic in the regional referral hospital in Makeni town during the time of the research. According to the result shown for the one hundred and twelve (112) participants interviewed, a huge percentage (83.0%) were not aware of their expected date of delivery, majority (79.5%) do not register for antenatal clinic within the first trimester of their pregnancy. Another huge percentage (72.3%) were unable to identify a choice of place of birth, with regards to identifying a skilled birth attendant (82.1%) were not able to identify a skilled birth attendant When it comes to doing shopping for birth supplies for delivery, the majority (70.5%) did not do shopping of essential items at the time of this study. Also, more than half of the participants (67.9%) did not know danger signs during pregnancy and a huge portion (84.8%) did not identify any means of transport to a health facility in case of any emergency. Slightly above half of the respondents (53.6%) had no one to accompany them to a health facility in time of labor and also majority of the participants (89.3%) at the time of study have no predetermined facility to access emergency services in case the need arises Close to the total number of participants (92.9%) had not got someone to donate blood for them if needed and some (50.89%) had not done savings in case an emergency at the time of the research. Some (62.5%) were not knowledgeable of childbirth and finally, (55.4%) were not aware of pregnancy complications.

**Table 3:** Level of Birth Preparedness and Knowledge Level in Pregnancy Danger Signs.

| **Variable** | **Frequency (n=112)** | **Percentage (%)** |
| --- | --- | --- |
| **Knowledge on expected date of delivery** |  |  |
| No | 93 | 83.04 |
| Yes | 19 | 16.96 |
| **Registered for ANC within the first trimester** |  |  |
| No | 89 | 79.46 |
| Yes | 23 | 20.54 |
| **Identification of choice for place of delivery** |  |  |
| No | 81 | 72.32 |
| Yes | 31 | 27.68 |
| **Identified any skilled birth attendant** |  |  |
| No | 92 | 82.14 |
| Yes | 20 | 17.86 |
| **Does your shopping for birth supplies for deliveries** |  |  |
| No | 79 | 70.54 |
| Yes | 33 | 29.46 |
| **Know of any danger sign during pregnancy** |  |  |
| No | 76 | 67.86 |
| Yes | 36 | 32.14 |
| **Have you identified a means of transport in case of emergency** |  |  |
| No | 95 | 84.82 |
| Yes | 17 | 15.18 |
| **Have you got a person to accompanying you during an emergency** |  |  |
| No | 60 | 53.57 |
| Yes | 52 | 46.43 |
| **Have you identified a predetermined place to access emergency service** |  |  |
| No | 100 | 89.29 |
| Yes | 12 | 10.71 |
| **Do you have someone to donate blood for you if needed** |  |  |
| No | 104 | 92.86 |
| Yes | 8 | 7.14 |
| **Have you been saving money in case of an emergency** |  |  |
| No | 57 | 50.89 |
| Yes | 55 | 49.11 |
| **Pre-knowledge on childbirth** |  |  |
| No | 70 | 62.5 |
| Yes | 42 | 37.5 |
| **Awareness on emergency complication** |  |  |
| No | 62 | 55.36 |
| Yes | 50 | 44.64 |

**Figure 1:**
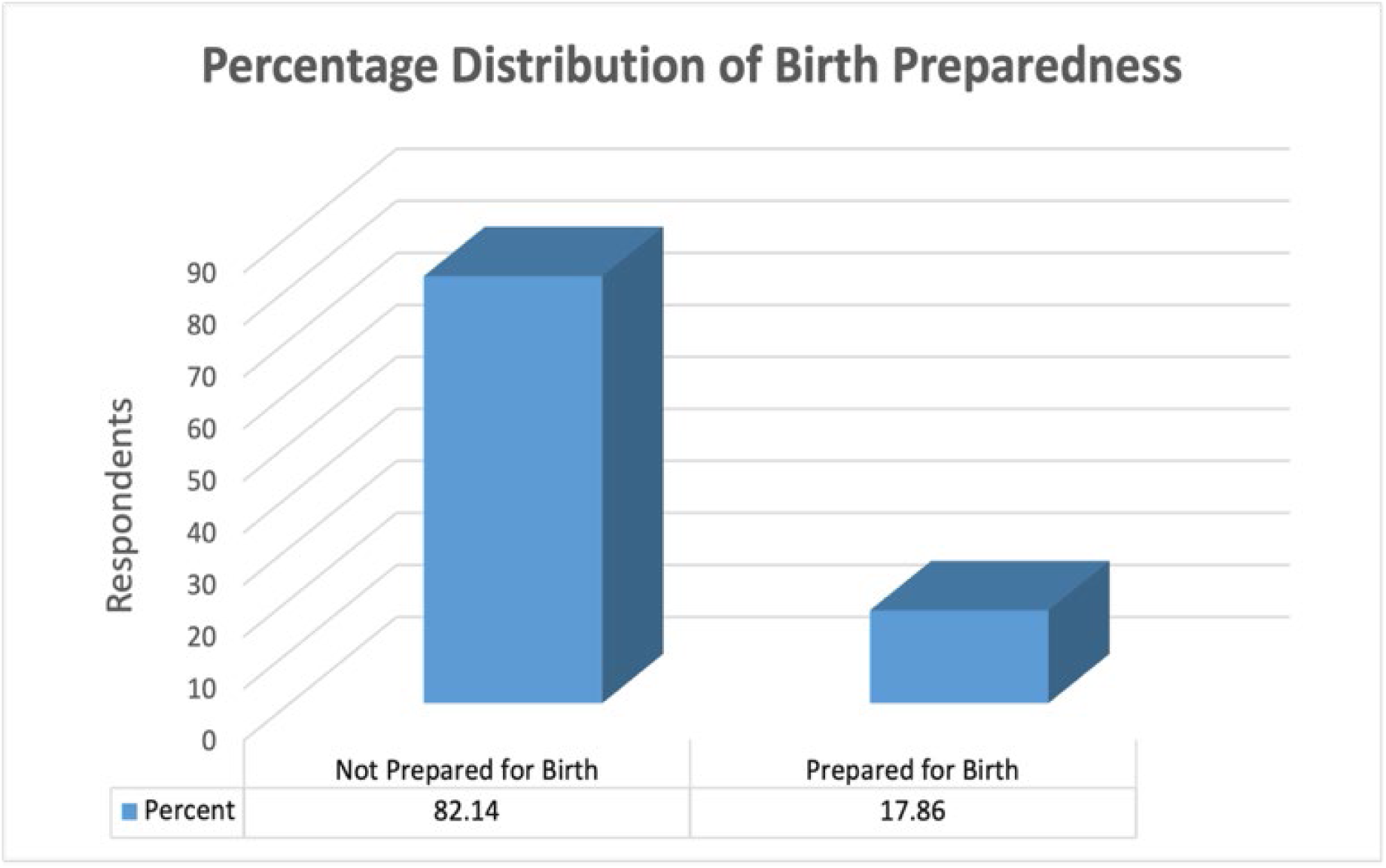
Percentage Distribution of Birth Preparedness

### 3.3 Economic Determinants of Birth Preparedness

The table below shows the economic determinants of both the respondents and their partners, in the table contains (2) portions. The top most part contains information relating to the sources of livelihood and monthly income of the participants and the lower part of the table contains information relating to the sources of livelihood and monthly earnings of the partners of the respondents. The sources of livelihood diverse ranging from business, some employed, others petty trading, some farmers and others get their livelihood from family and friends.

From the table above, 17 of the respondents which represents (15.18%) earns below 500Nle per month. 3 of the respondents earn within 501-1000Nle a month. Another 17 respondents accounting for (15.18%) earns on a monthly bases between 1001-1500Nle while the majority 74 of the participants accounting for (66.07%) gets a monthly income of between 1501-2000Nle and one of the respondents representing (0.89%) receives a monthly income above 2000Nle. With regards to the sources of livelihood of the partners of the respondents, some are bike riders, some business men, carpenters, drivers, farmers and lecturers. The result shows that (18. 75%) which is equal to 21 respondent’s partners earns 500Nle and below, (8.93%) which represent 10 partners earns between 501-1000Nle monthly. Also, (23.21%) which represent 26 respondent’s partners earn a monthly income between 1001-1500Nle while some 48 partners of participants accounting for (42.86%) receives a monthly bases 1501-2000Nle and the remaining 7 partners of the respondents which account for (6.25%) receive a monthly income above 2000Nle.

**Table 4:**
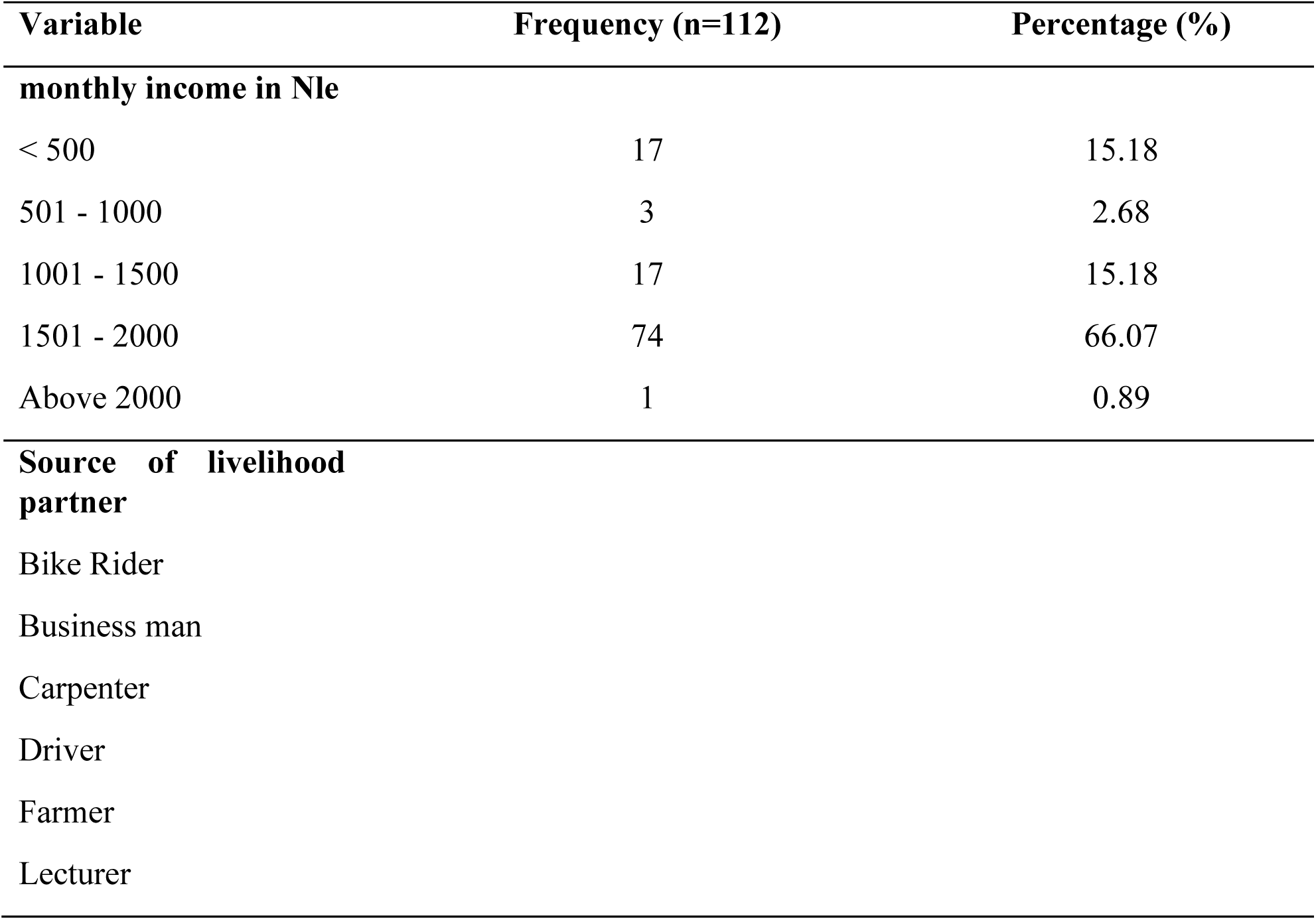
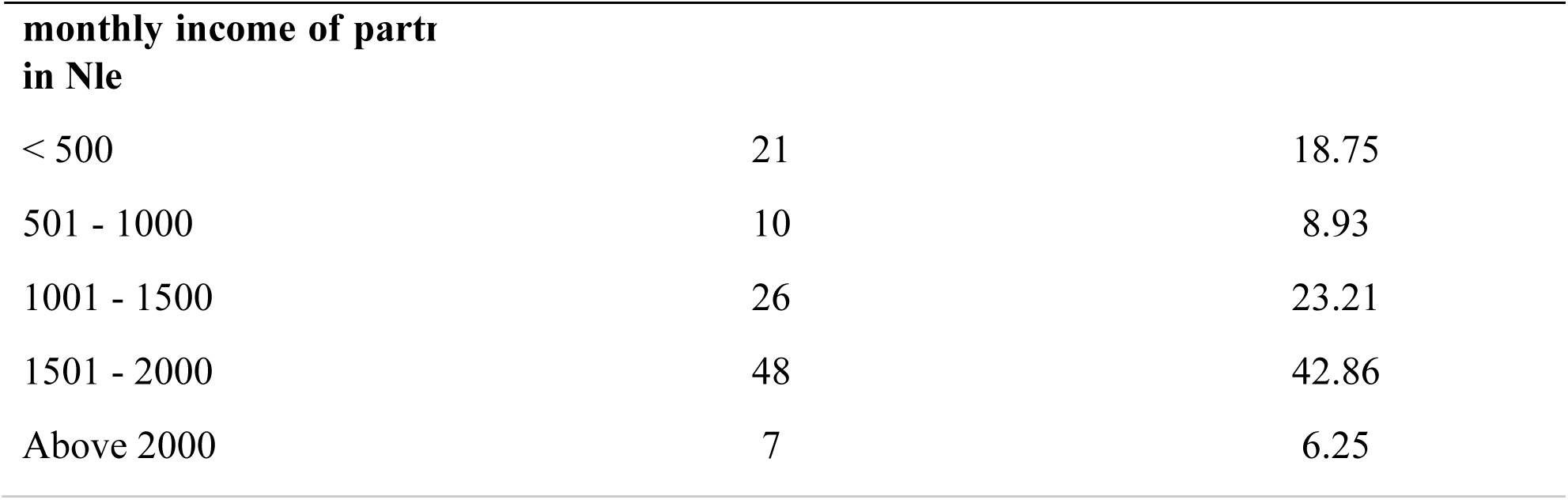
Economic determinants of Birth Preparedness.

### 3.4. Cultural and Behavioral Determinants of Birth Preparedness

The Table below describes the cultural and behavioral determinants of respondents. The questions were designed to determine the level of effect caused by cultural and behavioral factors on birth preparedness. A question was asked about food that cannot be consumed during pregnancy, a total number of one hundred and twelve (112) pregnant women were asked as respondents, and 22 of them which represent 19.64% said there are no foods that cannot be consumed by pregnant women however, 90 of the participants which represent 80.36% said there are foods that cannot be consumed by pregnant women. The questionnaire then asked about drinking alcohol during pregnancy, 70 of the respondents which represent 62.5% said that they don’t drink alcohol during pregnancy however, 42 of the respondents that represent 37.5% said yes they do drink alcohol during pregnancy. The questionnaire went further to ask about smoking cigarette during pregnancy, 56 of the respondents which represent 58.04% says no to smoking during pregnancy however, 47 participants which represents 41.96% said yes to smoking during pregnancy. When it comes to the level of awareness being raised in relation to substance abuse, 93 of the participants which represent (83.04%) of the respondents were not aware of the risk of premature birth as a result of substance abuse however, 19 of the participants were aware which represents (16.96%). The questionnaire went further to probe in to know whether the participants have any idea of the negative effect of substance abuse on the growth and development of the fetus; 101 of the participants said no which represent (90.18%) and 11 of them said yes which represent (9.82%). 36 of the women accounting for (32.14%) preferred to be delivered at home while the remaining 76 which represent (67.86%) preferred to be delivered in hospital. Of the 112 participants, 70 of them which accounts for (62.5%) preferred orthodox medicines while the remaining 42 which represent (37.5%) give preference to traditional medicines.

**Table 5:** Cultural and Behavioral Determinants of Birth Preparedness.

| **Variable** | **Frequency (n=112)** | **Percentage (%)** |
| --- | --- | --- |
| **Are there foods that you cannot eat while pregnant** |  |  |
| No | 22 | 19.64 |
| Yes | 90 | 80.36 |
| **Do you drink alcohol during pregnancy** |  |  |
| No | 70 | 62.5 |
| Yes | 42 | 37.5 |
| **Do you smoke cigarette during pregnancy** |  |  |
| No | 65 | 58.04 |
| Yes | 47 | 41.96 |
| **Awareness that substance abuse increases the risk of premature birth** |  |  |
| No | 93 | 83.04 |
| Yes | 19 | 16.96 |
| **Do you also know that substance abuse slows the growth and development of the fetus** |  |  |
| No | 101 | 90.18 |
| Yes | 11 | 9.82 |
| **Preferred place to delivered** |  |  |
| Home | 36 | 32.14 |
| Hospital | 76 | 67.86 |
| **preference of traditional medicine to orthodox medicine** |  |  |
| No | 70 | 62.5 |
| Yes | 42 | 37.5 |

### 3.5. Regression Analysis for Factors Influencing Birth Preparedness

From the regression analysis below to assess the determinants associated with birth preparedness of all demographic variables, the finding shows that the age of respondent [95% conf. interval = (-0.0006993-0.1082811), P-value = 0.05], and level of education [95% conf. interval = (0.0432869-0.1748262) P-value = 0.001] of respondents are significantly associated with birth preparedness. For instance, respondents within the age bracket 26-30 years are more likely to be involved in birth preparedness. This may imply that maybe those who are within age 26-30 have got experience through previous deliveries and have realized that childbirth preparation begins even before becoming pregnant. In some cases, partners support the pregnant women on every step of birth preparedness which has led to an increase number of the respondents. Talking of the other significant factor, which is the level of education of the respondents, this may suggest that respondents that are educated may have been thought in their institute of learning certain subject that has some bearing with childbirth for example, reproductive health, maternal and child health, to name but a few, which may have giving them some level of awareness. In some cases, women that are educated can be gainfully employed and due to their earnings they have the liberty to take decisions relating to childbirth with or without the approval of their partners. In the case of traditional practices regarding childbirth, respondents that are educated may not identify danger signs and relate it to tradition but they may go straight to a health facility and seek the service of a skilled service provider.

**Table 6:**
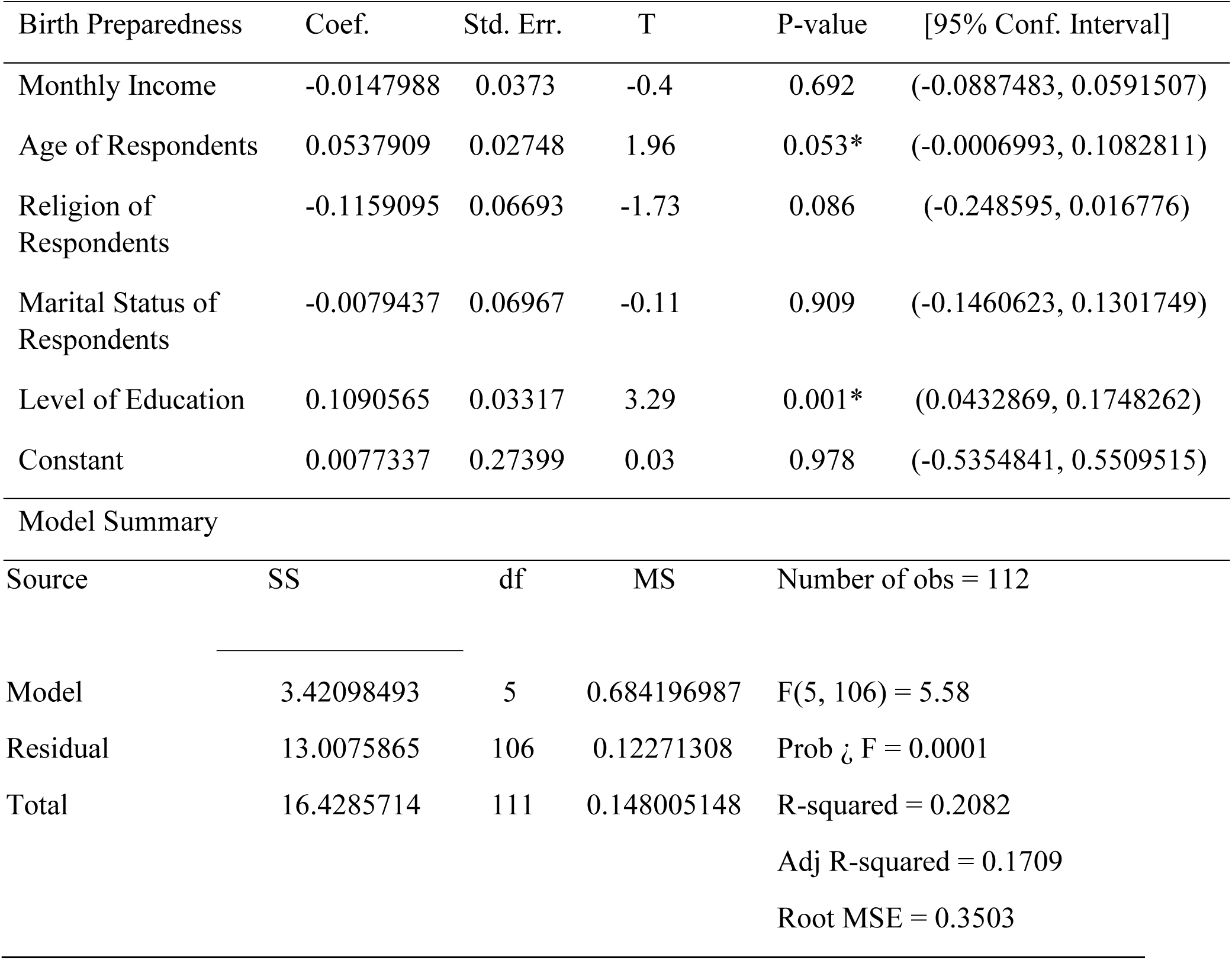
Factors Influencing Birth Preparedness.

| Birth Preparedness | Coef. | Std. Err. | T | P-value | [95% Conf. Interval] |
| --- | --- | --- | --- | --- | --- |
| Monthly Income | -0.0147988 | 0.0373 | -0.4 | 0.692 | (-0.0887483, 0.0591507) |
| Age of Respondents | 0.0537909 | 0.02748 | 1.96 | 0.053* | (-0.0006993, 0.1082811) |
| Religion of Respondents | -0.1159095 | 0.06693 | -1.73 | 0.086 | (-0.248595, 0.016776) |
| Marital Status of Respondents | -0.0079437 | 0.06967 | -0.11 | 0.909 | (-0.1460623, 0.1301749) |
| Level of Education | 0.1090565 | 0.03317 | 3.29 | 0.001* | (0.0432869, 0.1748262) |
| Constant | 0.0077337 | 0.27399 | 0.03 | 0.978 | (-0.5354841, 0.5509515) |
| Model Summary |  |  |  |  |  |
| Source | SS | df | MS | Number of obs = 112 |  |
| Model | 3.42098493 | 5 | 0.684196987 | F(5, 106) = 5.58 |  |
| Residual | 13.0075865 | 106 | 0.12271308 | Prob > F = 0.0001 |  |
| Total | 16.4285714 | 111 | 0.148005148 | R-squared = 0.2082 |  |
|  |  |  |  | Adj R-squared = 0.1709 |  |
|  |  |  |  | Root MSE = 0.3503 |  |

### 3.6. Logistic Regression of Socio-Demographic for Birth Preparedness

The logistic regression table shows the result of birth preparedness of respondents on the socio-demographic level, this table shows the significance of birth preparedness within the categories. In this situation, the age of respondents and the level of education of respondents are well explained here. In this table, the reference points are giving above. In the age category, the reference point is 20 years and below whereas in the area of education, the reference point is nonformal education. According to the result respondents that fall within age 26-30 years with an odd ratio = 10.27463 and a 95% confidence interval are more likely to prepare for childbirth compared to those within the ages of 20 and below years. Coming to the level education of respondents, it can be reported that respondents who attained a tertiary education with an odd ratio = 21.70098 and a (2.352484 - 200.1851) conf. Interval are 21 times likely to practice birth preparedness compared to those with non-formal education.

**Table 7:**
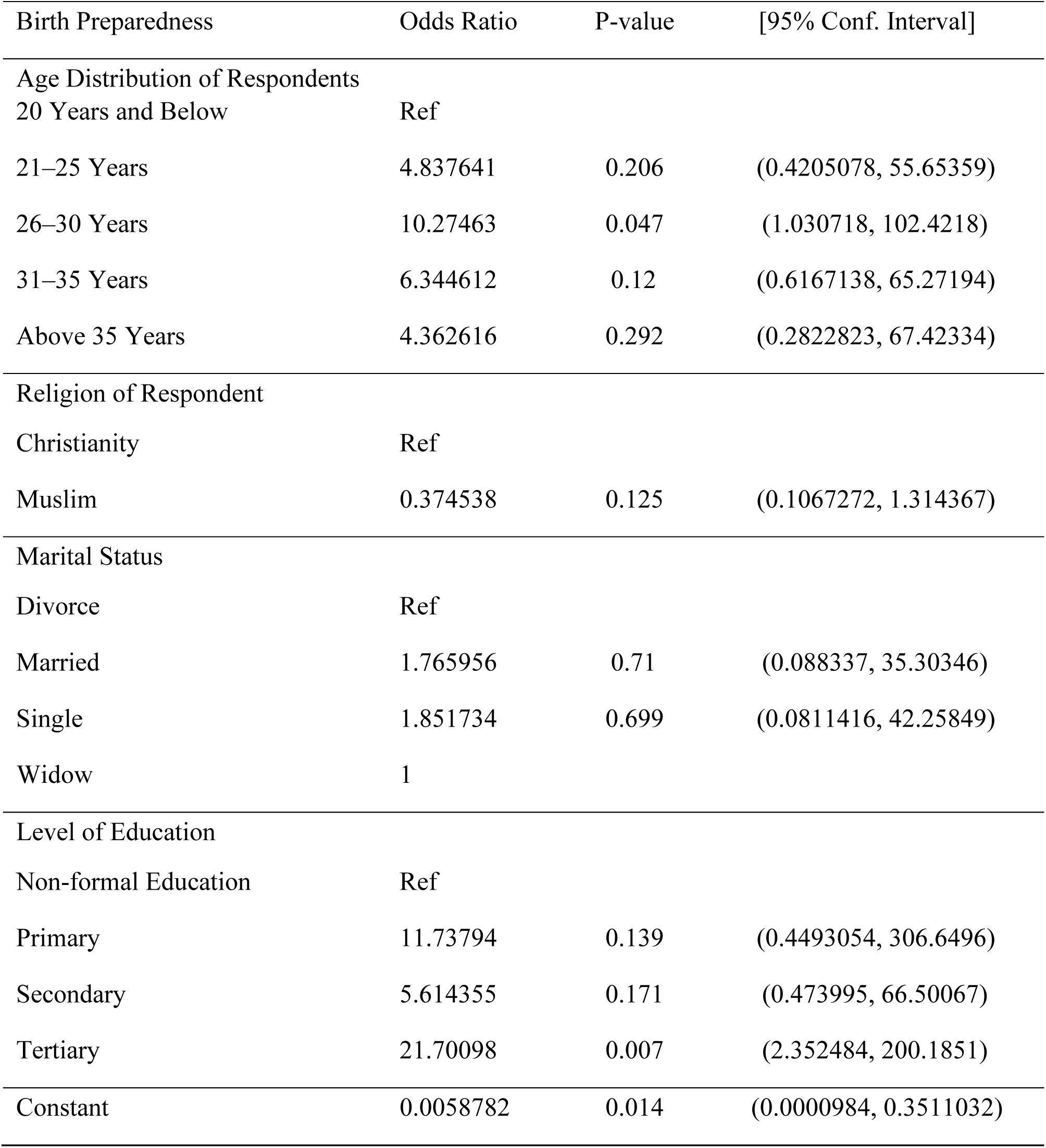
Logistic Regression of Socio-demographic for Birth Preparedness.

| Birth Preparedness | Odds Ratio | P-value | [95% Conf. Interval] |
| --- | --- | --- | --- |
| Age Distribution of Respondents |  |  |  |
| 20 Years and Below | Ref |  |  |
| 21–25 Years | 4.837641 | 0.206 | (0.4205078, 55.65359) |
| 26–30 Years | 10.27463 | 0.047 | (1.030718, 102.4218) |
| 31–35 Years | 6.344612 | 0.12 | (0.6167138, 65.27194) |
| Above 35 Years | 4.362616 | 0.292 | (0.2822823, 67.42334) |
| Religion of Respondent |  |  |  |
| Christianity | Ref |  |  |
| Muslim | 0.374538 | 0.125 | (0.1067272, 1.314367) |
| Marital Status |  |  |  |
| Divorce | Ref |  |  |
| Married | 1.765956 | 0.71 | (0.088337, 35.30346) |
| Single | 1.851734 | 0.699 | (0.0811416, 42.25849) |
| Widow | 1 |  |  |
| Level of Education |  |  |  |
| Non-formal Education | Ref |  |  |
| Primary | 11.73794 | 0.139 | (0.4493054, 306.6496) |
| Secondary | 5.614355 | 0.171 | (0.473995, 66.50067) |
| Tertiary | 21.70098 | 0.007 | (2.352484, 200.1851) |
| Constant | 0.0058782 | 0.014 | (0.0000984, 0.3511032) |

### 3.7 Age Distribution of Respondents by Birth Preparedness

In the chat ?, the analysis is based on the age distribution of respondents by birth preparedness of pregnant women attending antenatal clinic (ANC) in the regional referral hospital in Makeni city. The analysis is carried out to be able to identify the age bracket that could be prepared for child birth however, it could be realized from the analysis that from age 20 and below through age 35 and above were less prepared for child birth. It is clearly shown that pregnant women within the age brackets of twenty (20) and below that is the first age bracket recorded the highest number of forty (40) that were not prepared with minute number that were prepared for child birth. The second group is those pregnant women that were attending antenatal clinic (ANC) which were captured are those within the age brackets of 21 – 25 years. Ten of them according to the figure below not prepared also when you compare the first age group with the second, it seems the preparedness rate of the two are the same. The third age bracket is those within the age of 26 – 30; it could be clearly seen that more than ten (10) of the respondents were not prepared however, this age group is seen to have an encouraging number of respondents that were prepared for childbirth. The forth age group of respondent are those within age 31 – 35 years, according to the figure below, ten (10) of the respondents were not prepared for childbirth and less than half (1/2) of ten (10) were prepared for childbirth and finally, the last age group is the ones above 35 years. According to the graph not up to ten (10) of the respondents that are above 35 years of age were prepared and only a minute number of them were found to be prepared for child. In summary, birth preparedness among pregnant women attending antenatal clinic in the regional referral hospital in Makeni city is found to be very low across all ages, the lack of birth preparedness practice prominent among young ladies that are within age twenty (20) and below.

**Figure 2:**
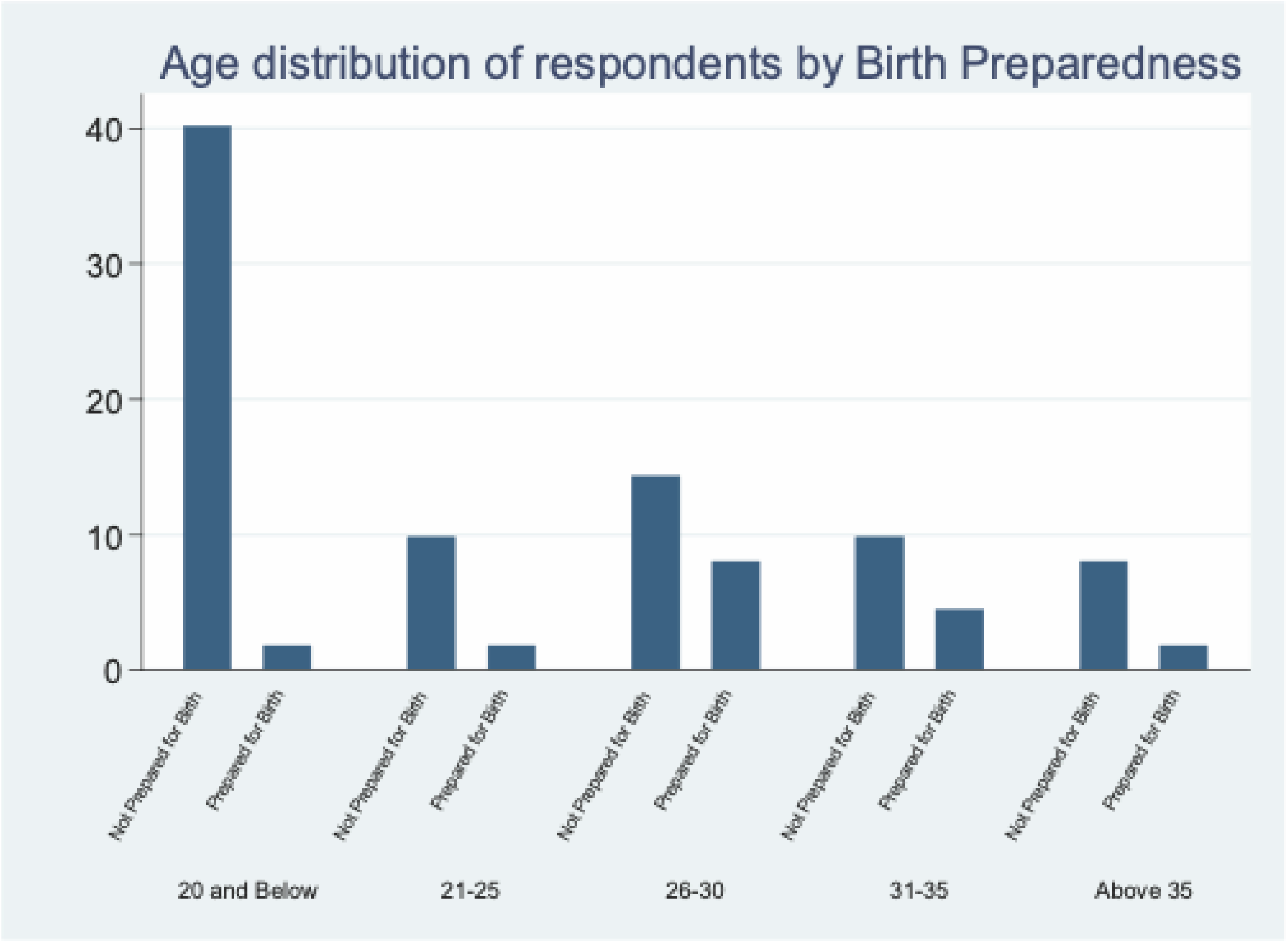
A**g**e **Distribution of Respondents by Birth Preparedness**

## 4. DISCUSSION

The study aimed to evaluate factors influencing birth preparedness among pregnant women at the Antenatal Care Clinics of Makeni Regional Referral Hospital. Birth preparedness involves strategies to ensure timely access to skilled birth attendants, awareness of pregnancy danger signs, arranging emergency transportation, securing a potential blood donor, shopping for childbirth supplies, selecting a delivery location, registering for antenatal care early, and understanding pregnancy complications. Knowledge of the expected delivery date is critical for safe motherhood, reducing maternal and neonatal mortality. Our research provides insights into the knowledge levels of pregnant women at the study facility, offering a foundation for designing targeted interventions and broader studies across other facilities in Sierra Lone. The findings highlight significant gaps in birth preparedness. Awareness of pregnancy danger signs was low, indicating a need for enhanced health education and community sensitization efforts. Early ANC registration, crucial for safe pregnancy and delivery, was also limited. Many respondents had not identified a delivery facility or a skilled birth attendant, potentially influenced by factors such as age and education level. Additionally, preparedness for emergencies was inadequate, with few participants having arranged transportation or identified a companion for emergencies. However, only half of the respondents had saved funds for emergencies and identified a health facility for emergency services. Awareness of pregnancy complications was moderate, and a small number had secured a blood donor or had prior childbirth knowledge. Regression and logistic regression analyses were conducted to identify significant variables affecting birth preparedness. The results indicated that the age of respondents and their education level were statistically significant factors. Specifically, younger age groups and those with higher education, particularly at the tertiary level, showed greater preparedness, possibly due to prior childbirth experience, awareness, sensitization, or partner involvement. Overall, the study reveals low levels of birth preparedness among participants, underscoring the need for targeted interventions to improve maternal and neonatal outcomes.

## 5. CONCLUSION

This study underscores the critical gaps in birth preparedness among pregnant women attending the Antenatal Care Clinics at Makeni Regional Referral Hospital. The low levels of awareness regarding pregnancy danger signs, limited early ANC registration, and inadequate emergency preparedness highlight the urgent need for enhanced maternal health interventions. The significant influence of age and education level on preparedness suggests that younger and more educated women are better equipped for childbirth, likely due to prior experience or access to information. These findings emphasize the importance of addressing disparities in knowledge and access to resources, particularly for less-educated and first-time mothers. A proposed policy intervention, including community-based education, improved access to skilled care, and emergency transportation networks, will offer a pathway to strengthen maternal healthcare systems. By implementing these strategies, health authorities can enhance birth preparedness, reduce maternal and neonatal mortality, and promote equitable health outcomes. Further research is recommended to explore these factors in other regions of Sierre Leone and to evaluate the effectiveness of targeted interventions in improving birth preparedness across diverse populations.

## STATEMENTS and DECLARATIONS

## ABBREVIATIONS

ANC: Antenatal Clinic Care
UNDP: United Nation Development Program
BP: Birth Preparedness
M10: Matturie Birth Preparedness Scale
UNIMAK: University of Makeni
MM: Maternal Mortality
MDGs: Millennium Development Goals
SSA: Sub-Sahara Africa
MMR: Maternal Mortality Rate

## Data Availability

All data produced in the present work are contained in the manuscript.

## ACKNOWLEDGEMENT

We thank the data collection team members for their exemplary work, the Sierra Leone Ministry of Health and Sanitation for giving access to their facility and data sources for this study, and the administration of the University of Makeni (UNIMAK) for the provision of students who helped with the management of the data collection process.

## DATA AVAILABILITY

All data for our research study are presented within this article and are available separately from the Lead Author or Corresponding Author with a written or electronic request from any interested individual.

## TRANSPARENCY

The authors volunteer that *chat GPT*, or any equivalent AI program, was not a source of data or information and was not used to draft or embellish our manuscript.

## ETHICAL APPROVAL & INFORMED CONSENT STATEMENT

Ethical approval was sought and received from the Ethical Review Board of the Sierra Leone Ethics and Scientific Review Committee. An inform consent form was issued to each respondent to seek their knowledge and to assure them that their contribution to the study will be confidential and the data provided can only be used for the purpose of the study. The respondents were also informed that no form of financial incentive will be given to them for being part of the study, and that their choice of not being part of the study cannot stop them from getting any benefit that may come because of the study.

## AUTHORS CONTRIBUTIONS

Tamba Matturie Conceptualization, Administration, Formal Data Analysis, Investigation, and Formalizing Original Manuscript.

Abraham Isiaka Jimmy Co-Conceptualization, Extra Data Analysis, and Writing – Drafting Original Manuscript and Reviewing & Editing. Tenneh Millicent Conteh Data Collection, Monitoring, and Co-Data Management. Alusine Rhoda Thullah Data Analysis, Validation, and Co-Drafting Manuscript. Magdalene Philip Umoh Supervision, Methodology, and Data Management.

Rebecca Esliker Supervision, Validation, and Co-Editing Manuscript.

Lee Presley Gary Jr. Resources, Visualization, and Writing – Restructuring,

Reviewing & Editing Final Manuscript.

## BIOGRAPHIES

**Abraham Isiaka Jimmy** is a Public Health Specialist and Researcher and University Instructor. He has more than six years of experience working in emergency management and community development contexts in Sierra Leone, while focusing on improving health program management. He teaches public health and mental health courses at the University of Makeni, located in the City of Maken, Sierra Leone. He advises on the operationalization of One Health Programs, including developing policies, strategies, and governance for mitigating public health challenges and identifying evidence-based solutions.

**Tenneh Millicent Conteh** is a self-motivated, result oriented and dedicated. She always subordinates personal interest to that of a group/organization; aims at excellence as an individual or in a team to seek solutions to challenging situations in life, a quick learner, flexible with a mind to learn, work and to impart, as well as good interpersonal and strong analytical skills. She is a collaborator with good organizational and management skills and can work with or without supervision. Proven track record to meet schedule and deliver high-quality results in both Academic and Non-governmental Organizations in Sierra Leone, with more than five years of experience in public health-related issues and community health improvement.

**Magdalene Philip Umoh** is a Public Health Scholar with expertise in Occupational and Environmental Health and Safety. She is a faculty lecturer for the Department of Public Health at the University of Makeni, located in Makeni, Sierra Leone, and serves as Director of the Center of Excellence in Maternal and Child Health Education and Research. Her vast experience with public and community health issues covers over ten years. including lecturing, researching, and consulting with the Food and Agricultural Organization (FAO), Ministry of Agriculture, and various Human Capital Development Plans (HCD Plus). She is passionate about transforming public health outcomes with a keen focus on women and children.

**Lee P. Gary, Jr.** is a Visiting Fulbright Research Scholar in the Department of Public Health at the University of Malta for 2025-2026. During 2024, he was a Visiting Research Scholar at the University of Makeni, located in the City of Makeni, Sierra Leone, and he is Owner/CEO of Strategic Management Services–USA, a global consultancy specializing in mitigating public health threats and hazards associated with untreated wastewater. He is a member of the Water Environment Federation and is an adjunct instructor for the National Disaster & Emergency Management University (Emmitsburg, Maryland, USA).

## RESEARCH FIELDS

**Abraham Isiaka Jimmy:** Public Health Policy, Health System Financing, Healthcare Economics, Public Mental Health Policy, Maternal Child Health, Occupational Health Safety & Regulations, and Environmental Health Risks & Assessments.

**Tenneh Millicent Conteh:** Public Health Management, Health Policy Issues Development & Strategy, Occupational Health Safety & Regulations, Environmental Health Challenges & Training, and Infectious Diseases Surveillance & Monitoring.

**Magdalene Philip Umoh:** Maternal Healthcare, Occupational Health Safety & Regulations, Environmental Health Risk & Assessments, Public Health Policy, Infectious Diseases, and Nursing Profession Education & Career Development.

**Rebecca Esliker:** Higher Education Leadership & Administration, Public Health Curriculum & Training, Communicable and Non-communicable Diseases, Population Health Awareness & Surveillance, and Disaster management.

**Lee Presley Gary, Jr.:** Disaster Management, Emergency Management, and Mitigation of Dirty Water and Human Waste, and Professional Development of Healthcare Workforce.

**\* * * * \***

